## Supplemental Information for "Wastewater surveillance reveals patterns of antibiotic resistance across the United States"

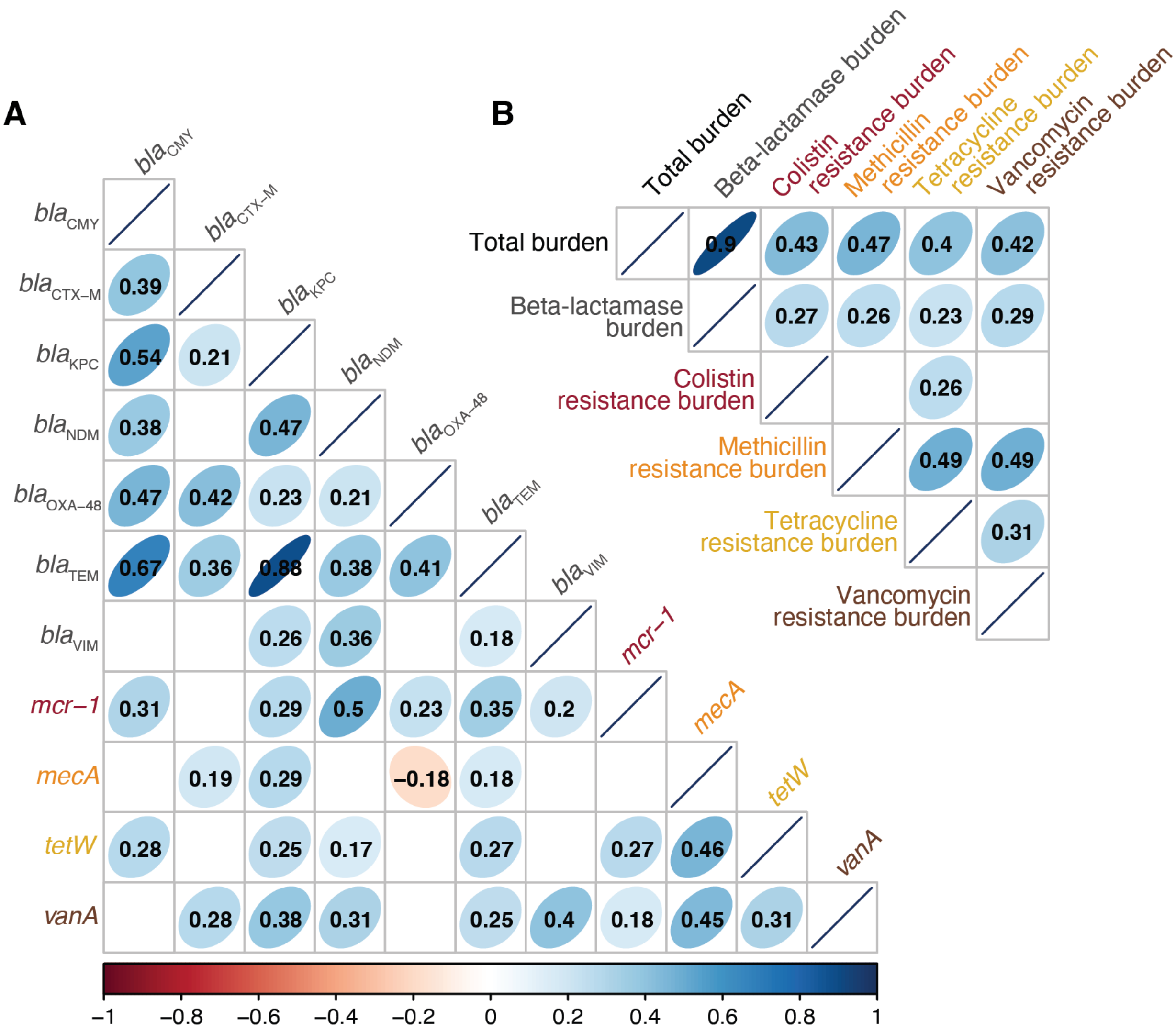


**Figure S1**. **Correlation among ARGs measured in wastewater. A)** Spearman's correlation coefficient among ARG concentrations normalized by 16S rRNA gene ($p < 0.05$). **B)** Spearman's correlation coefficient among antibiotic resistance burden calculated as z-scores based on distribution of each gene ($p < 0.05$).

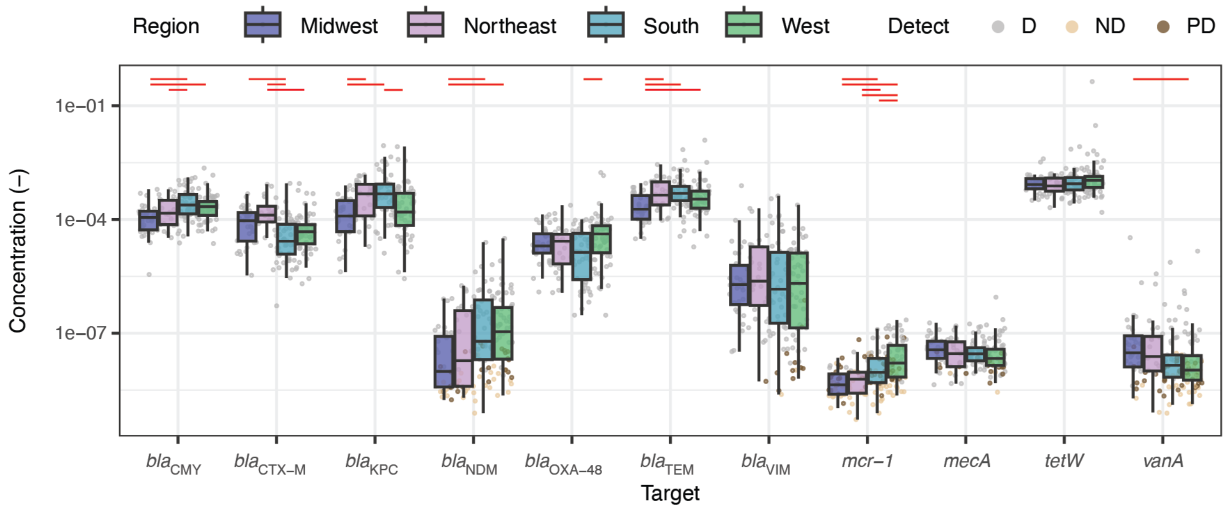


**Figure S2. Regional distribution of all ARGs.** Each data point represents a wastewater treatment plant. Samples were categorized as non-detect (ND) if none of the replicates detected the target, detect (D) if all replicates detected it, and partial detect (PD) if at least one of the replicates did not detect the target. If the gene was undetected, half of the theoretical measurement limit was substituted as the measured value. The median is shown by the line inside the box with the 25th and 75th percentile represented by the lower and upper boundary of the box. Bottom and top whiskers show 1.5 x interquartile range. Red lines indicate a significant pairwise difference between regions measured by the two-tail Conover-Iman post-hoc test with Benjamini-Hochberg correction applied ($p < 0.025$).

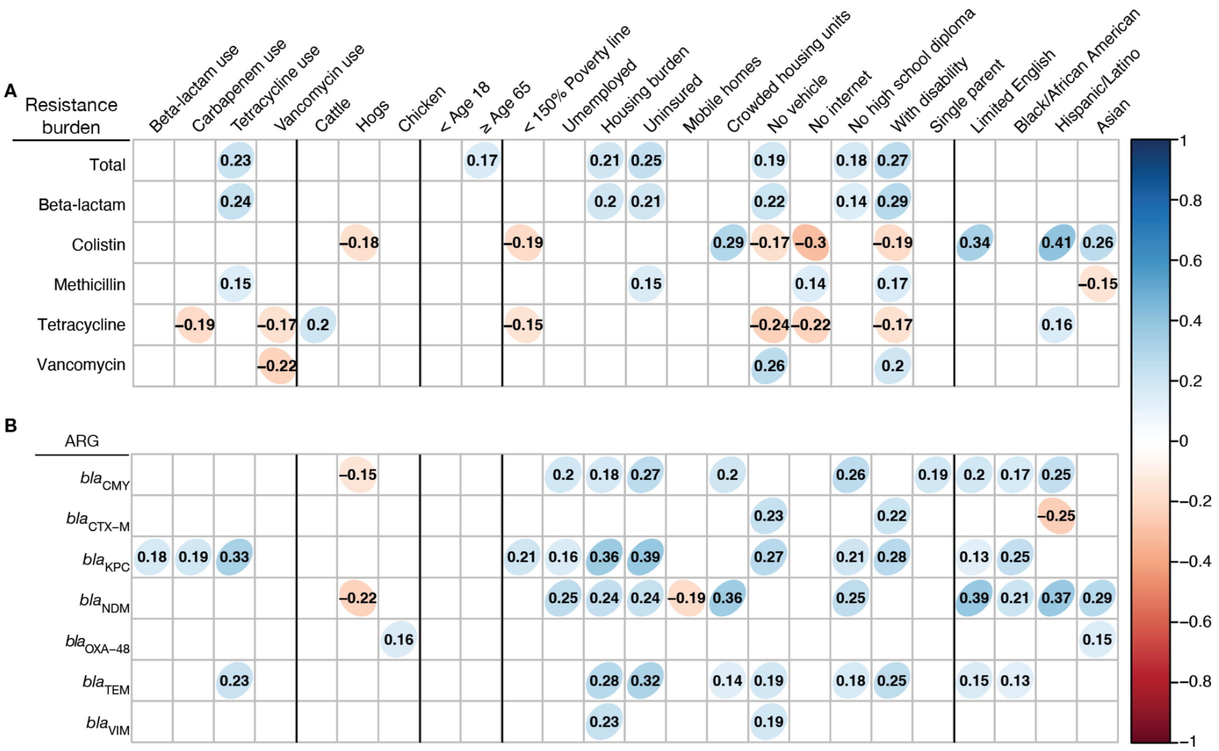

**Figure S3. Unadjusted correlation resistance measured in wastewater and potential determinants.** Spearman correlation coefficient among potential determinants of resistance burden and **A)** antibiotic resistance burden score and **B)** beta-lactamase gene concentrations measured in wastewater and normalized by 16S rRNA gene ($p < 0.05$).

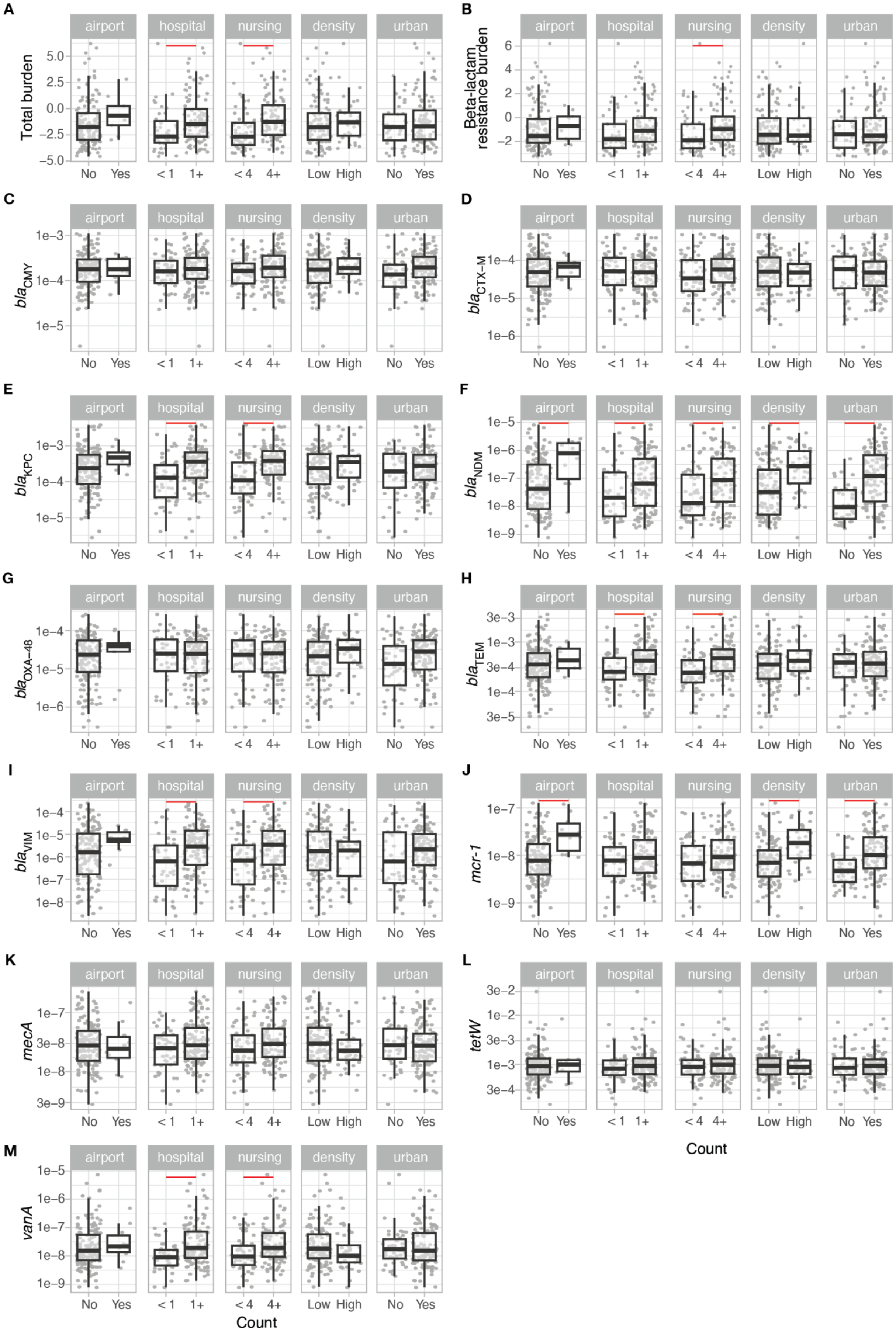
A
B
C
D
E
F
G
H
I
J
K
L
M
airport
hospital
nursing
density
urban
Total burden
Beta-lactam resistance burden
$bla_{CMY}$
$bla_{CTX-M}$
$bla_{KPC}$
$bla_{NDM}$
$bla_{OXA-48}$
$bla_{TEM}$
$bla_{VIM}$
mcr-1
mecA
tetW
vanA
No
Yes
< 1
1+
< 4
4+
Low
High
Count

**Figure S4. Comparison of categorical variables for antibiotic resistance burden and genes**. Bivariate analysis of points of interest, population density, and urbanicity for **A)** total resistance burden and **B)** beta-lactamase resistance burden measured in wastewater as aggregate z-scores. Same analysis done for wastewater concentration normalized by 16S rRNA gene for individual genes, **C)** $bla_{\text{CMY}}$, **D)** $bla_{\text{CTX-M}}$, **E)** $bla_{\text{KPC}}$, **F)** $bla_{\text{NDM}}$, **G)** $bla_{\text{OXA-48}}$, **H)** $bla_{\text{TEM}}$, **I)** $bla_{\text{VIM}}$, **J)** *mcr-1*, **K)** *mecA*, **L)** *tetW*, and **M)** *vanA*. The median is shown by the line inside the box with the 25th and 75th percentile represented by the lower and upper boundary of the box. Bottom and top whiskers show 1.5 x interquartile range. Red lines indicate a significant pairwise difference between regions measured by the Wilcoxon post-hoc test with Benjamini-Hochberg correction applied ($p < 0.05$).

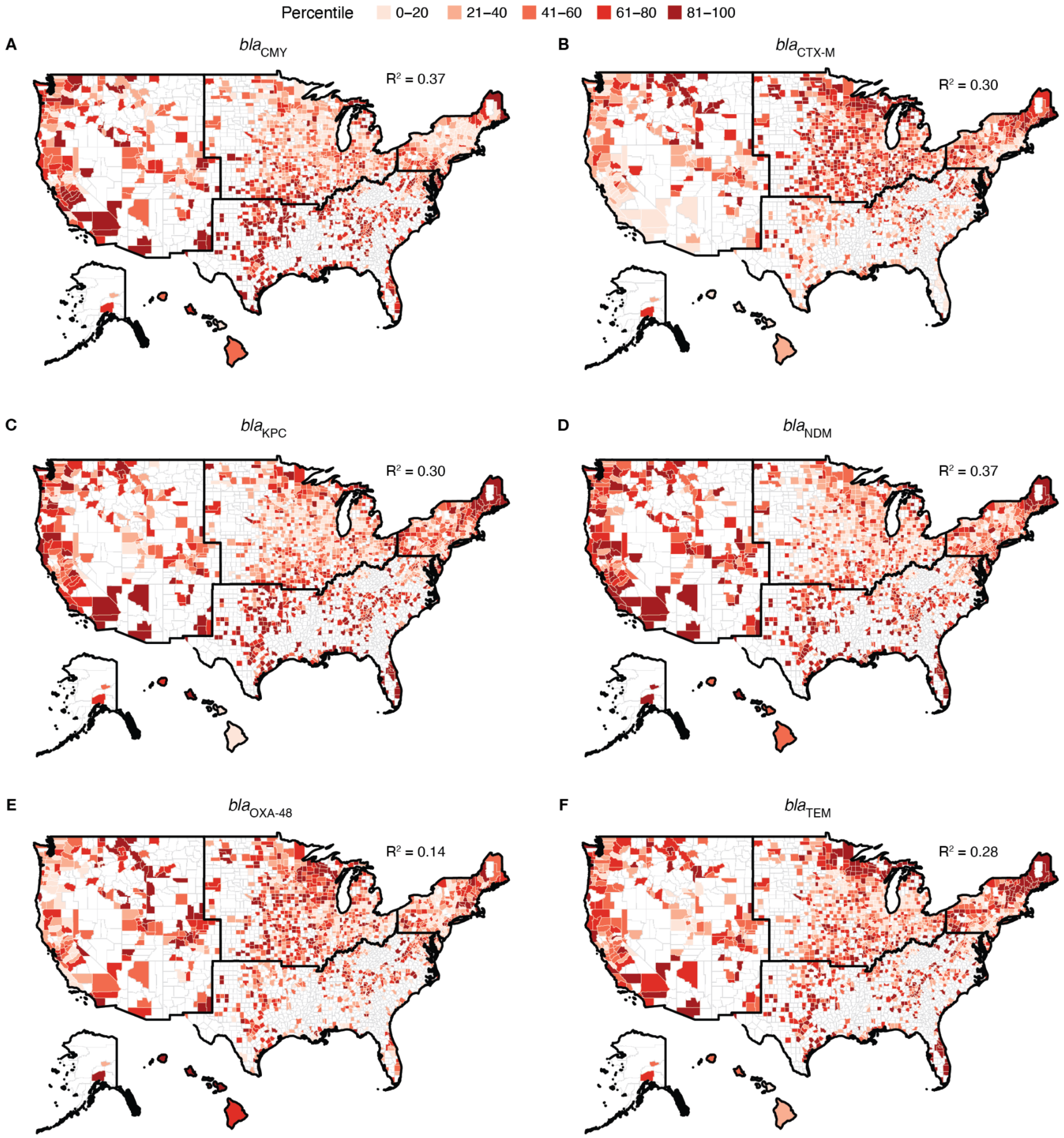
Percentile
0–20
21–40
41–60
61–80
81–100
A
bla CMY
R² = 0.37
B
bla CTX-M
R² = 0.30
C
bla KPC
R² = 0.30
D
bla NDM
R² = 0.37
E
bla OXA-48
R² = 0.14
F
bla TEM
R² = 0.28

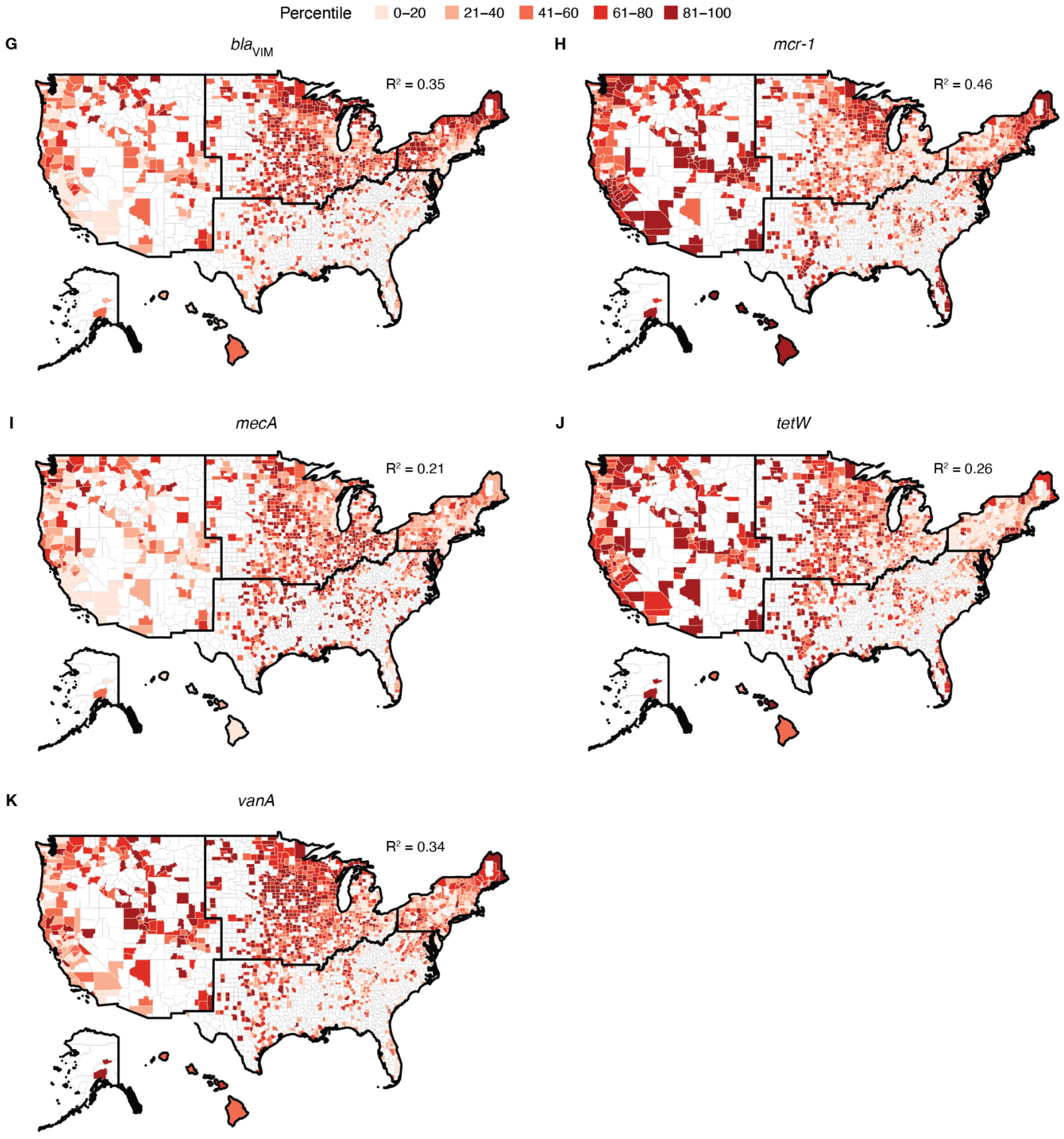


**Figure S5. Random forest modeling of antibiotic resistance gene concentrations across the United States using secondary data.** Predicted **A)** *bla*$_{CMY}$ ($R^2$: 0.37), **B)** *bla*$_{CTX-M}$ ($R^2$: 0.30), **C)** *bla*$_{KPC}$ ($R^2$: 0.30), **D)** *bla*$_{NDM}$ (R2: 0.37), **E)** *bla*$_{OXA-48}$ ($R^2$: 0.14), **F)** *bla*$_{TEM}$ ($R^2$: 0.28), **G)** *bla*$_{VIM}$ ($R^2$: 0.35), **H)** *mcr-1* ($R^2$: 0.46), **I)** *mecA* ($R^2$: 0.21), **J)** *tetW* ($R^2$: 0.26), and **K)** *vanA* ($R^2$: 0.34) concentrations across the United States. The predicted concentration, normalized by 16S rRNA, is visualized as percentiles, with darker colors indicating higher concentrations. The random forest models are trained based on the secondary data in Table 1. A higher percentile indicates a higher predicted concentration within that county. Each map shows predictive performance of the model indicated by $R^2$. Alaska and Hawaii are not to scale.

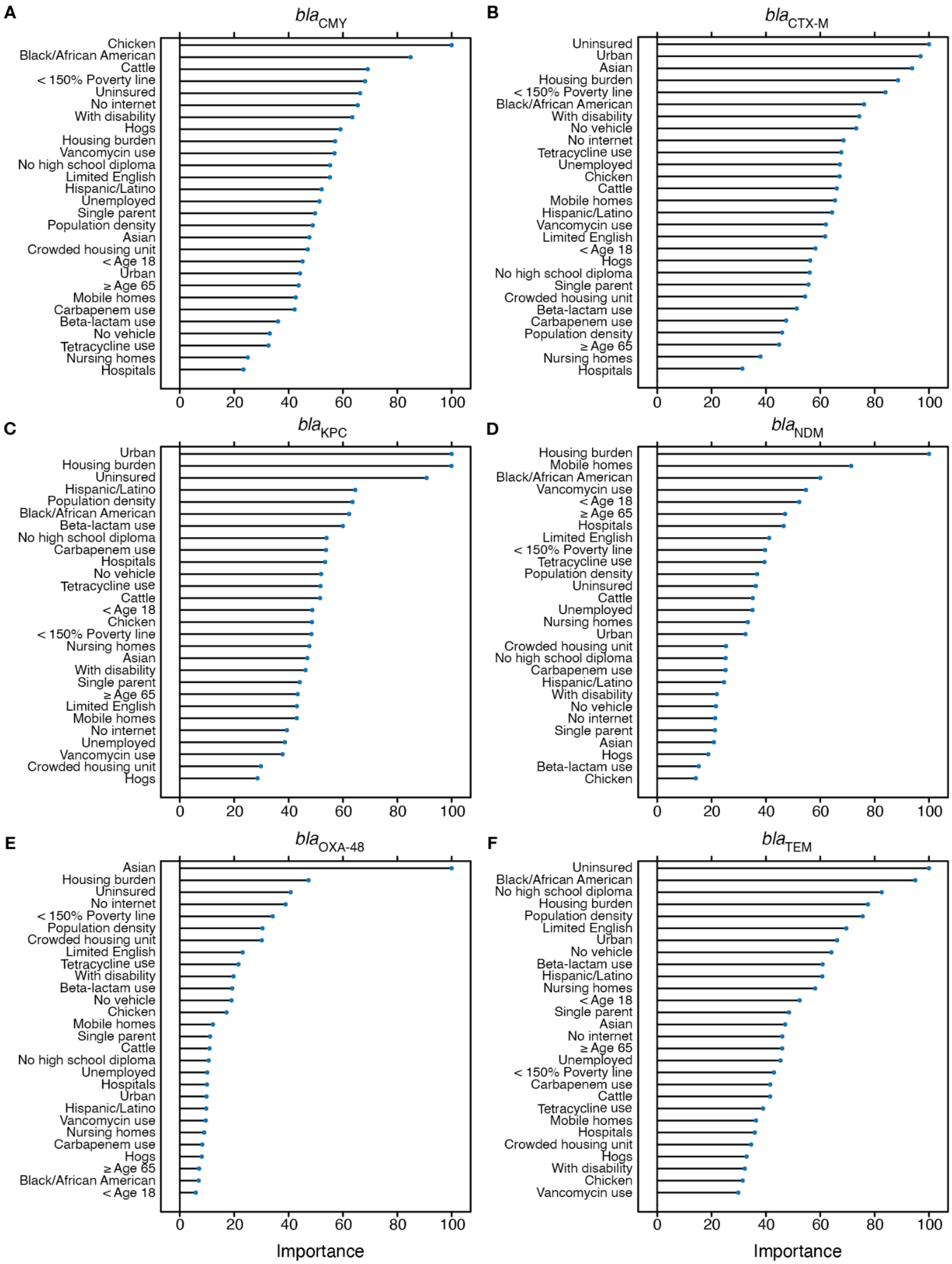
A
$bla_{CMY}$
Chicken
Black/African American
Cattle
< 150% Poverty line
Uninsured
No internet
With disability
Hogs
Housing burden
Vancomycin use
No high school diploma
Limited English
Hispanic/Latino
Unemployed
Single parent
Population density
Asian
Crowded housing unit
< Age 18
Urban
≥ Age 65
Mobile homes
Carbapenem use
Beta-lactam use
No vehicle
Tetracycline use
Nursing homes
Hospitals
B
$bla_{CTX-M}$
Uninsured
Urban
Asian
Housing burden
< 150% Poverty line
Black/African American
With disability
No vehicle
No internet
Tetracycline use
Unemployed
Chicken
Cattle
Mobile homes
Hispanic/Latino
Vancomycin use
Limited English
< Age 18
Hogs
No high school diploma
Single parent
Crowded housing unit
Beta-lactam use
Carbapenem use
Population density
≥ Age 65
Nursing homes
Hospitals
C
$bla_{KPC}$
Urban
Housing burden
Uninsured
Hispanic/Latino
Population density
Black/African American
Beta-lactam use
No high school diploma
Carbapenem use
Hospitals
No vehicle
Tetracycline use
Cattle
< Age 18
Chicken
< 150% Poverty line
Nursing homes
Asian
With disability
Single parent
≥ Age 65
Limited English
Mobile homes
No internet
Unemployed
Vancomycin use
Crowded housing unit
Hogs
D
$bla_{NDM}$
Housing burden
Mobile homes
Black/African American
Vancomycin use
< Age 18
≥ Age 65
Hospitals
Limited English
< 150% Poverty line
Tetracycline use
Population density
Uninsured
Cattle
Unemployed
Nursing homes
Urban
Crowded housing unit
No high school diploma
Carbapenem use
Hispanic/Latino
With disability
No vehicle
No internet
Single parent
Asian
Hogs
Beta-lactam use
Chicken
E
$bla_{OXA-48}$
Asian
Housing burden
Uninsured
No internet
< 150% Poverty line
Population density
Crowded housing unit
Limited English
Tetracycline use
With disability
Beta-lactam use
No vehicle
Chicken
Mobile homes
Single parent
Cattle
No high school diploma
Unemployed
Hospitals
Urban
Hispanic/Latino
Vancomycin use
Nursing homes
Carbapenem use
Hogs
≥ Age 65
Black/African American
< Age 18
F
$bla_{TEM}$
Uninsured
Black/African American
No high school diploma
Housing burden
Population density
Limited English
Urban
No vehicle
Beta-lactam use
Hispanic/Latino
Nursing homes
< Age 18
Single parent
Asian
No internet
≥ Age 65
Unemployed
< 150% Poverty line
Carbapenem use
Cattle
Tetracycline use
Mobile homes
Hospitals
Crowded housing unit
Hogs
With disability
Chicken
Vancomycin use
0
20
40
60
80
100
Importance

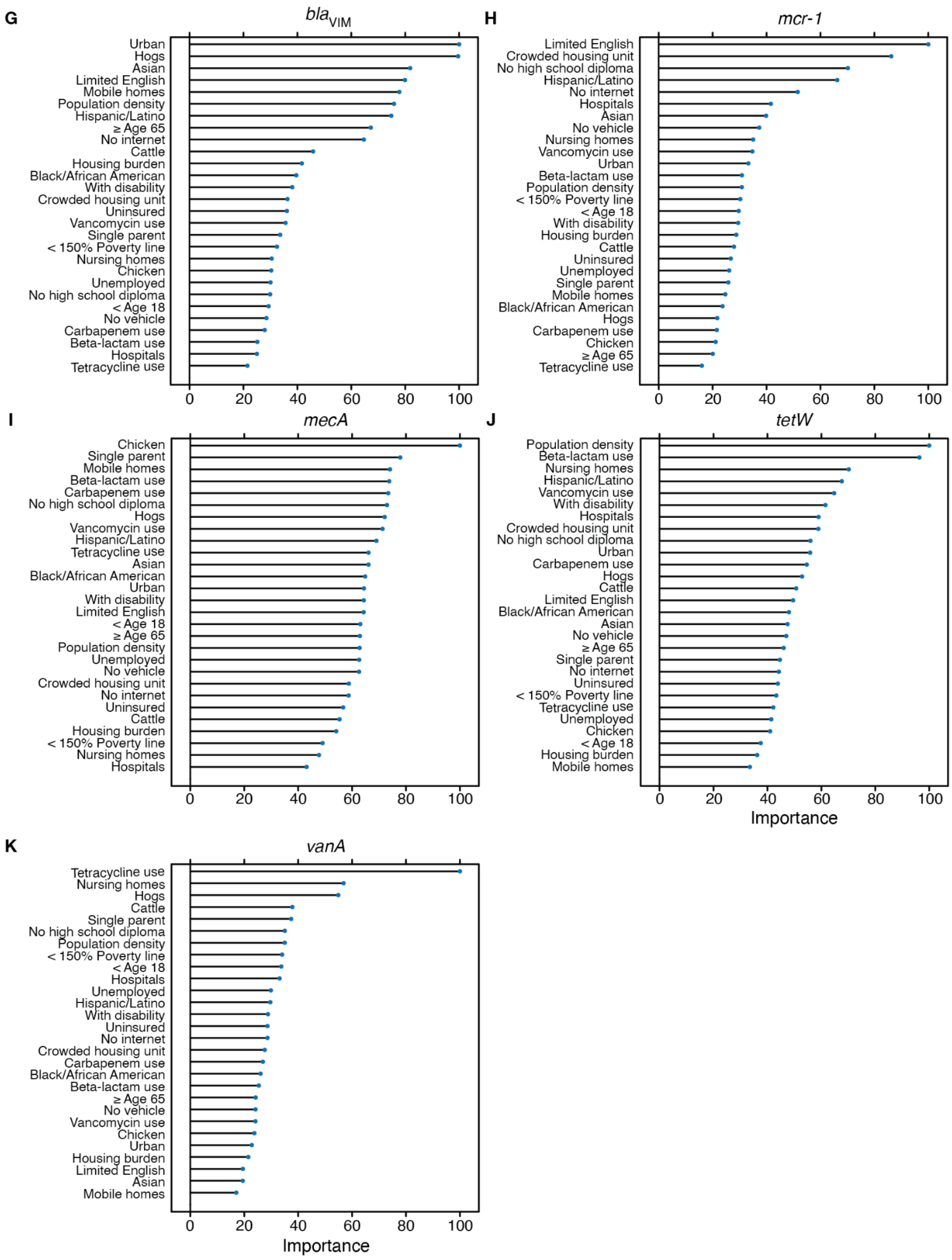


**Figure S6. Variable importance for individual ARG random forest prediction model.** Random forest model predicts relative concentration of ARGs normalized by 16S rRNA across all counties in the U.S. for **A)** *bla*$_{CMY}$, **B)** *bla*$_{CTX\text{-}M}$, **C)** *bla*$_{KPC}$, **D)** *bla*$_{NDM}$, **E)** *bla*$_{OXA\text{-}48}$, **F)** *bla*$_{TEM}$, **G)** *bla*$_{VIM}$, **H)** *mcr-1*, **I)** *mecA*, **J)** *tetW*, and **K)** *vanA*.

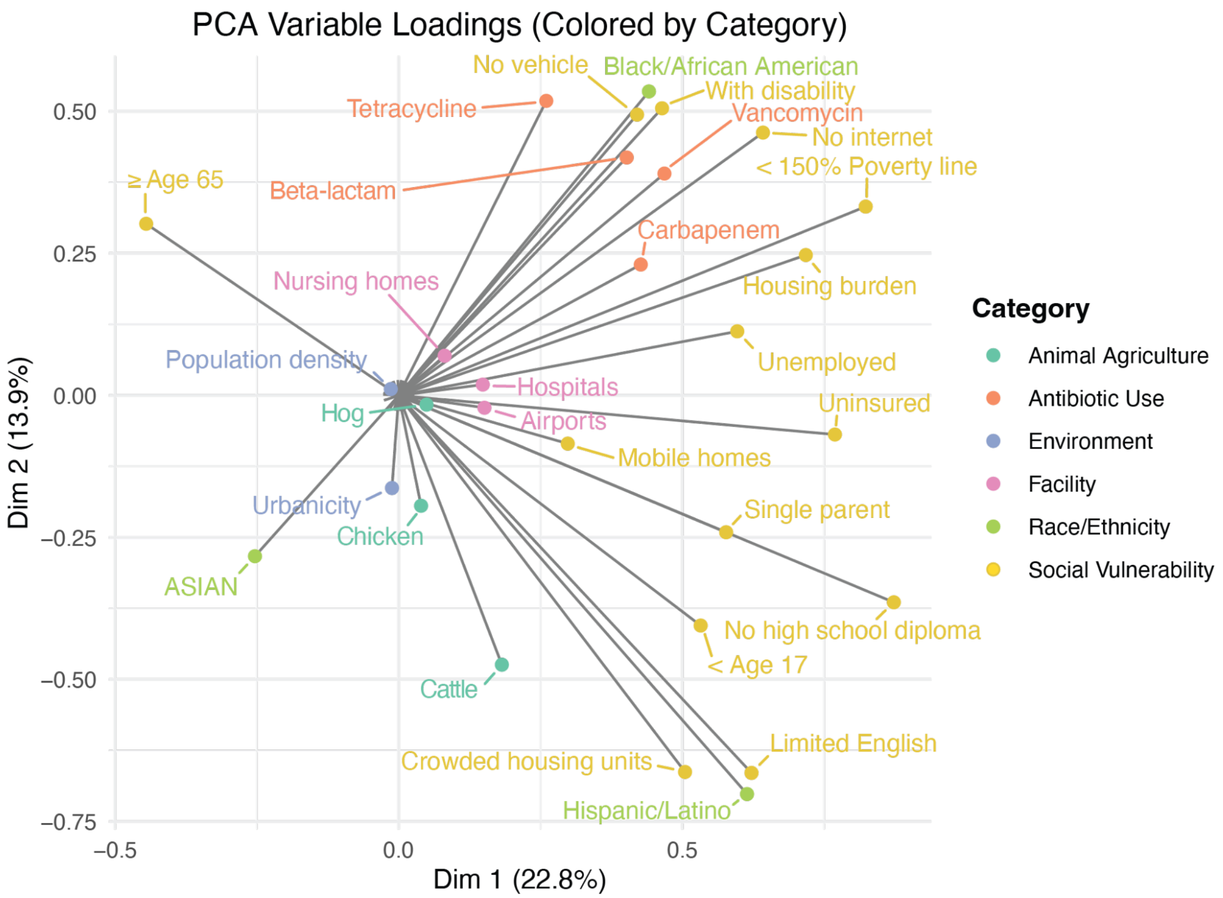


**Figure S7. Principle component analysis (PCA) of secondary variables used in the study.** A Principal Component Analysis (PCA) was performed on the metadata parameters to investigate the underlying structure and potential co-correlation. Secondary variables are categorized by color. The PCA indicated that the variance within the metadata was distributed across multiple dimensions, with the first two principal components explaining 36.7% of the total variance. Visual inspection of variable loadings revealed that while the social vulnerability indicators generally loaded positively on Dimension 1, suggesting a shared underlying characteristic, they displayed a considerable spread among Dimension 2. Subsequent components (PC3: 11.5%, PC4: 8.7%) captured further, distinct aspects of the data structure.

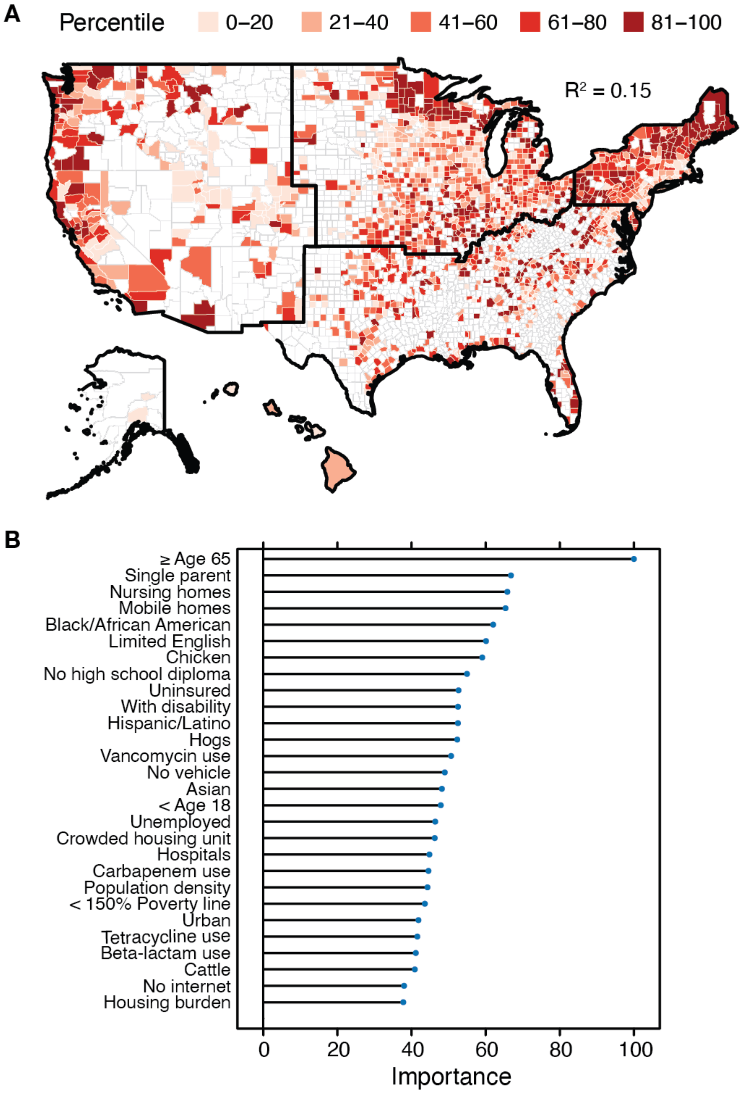


**Figure S8. Random forest modeling of overall antibiotic resistance gene prevalence across the United States using secondary data. A)** The predicted resistance gene prevalence and **B)** variable importance using cumulative burden z-scores visualized as percentiles, with darker colors indicating higher concentrations. The random forest models are trained based on the secondary data in Table 1. A higher percentile indicates a higher predicted concentration within that county. Predictive performance of the model indicated by $R^2$. Alaska and Hawaii are not to scale.

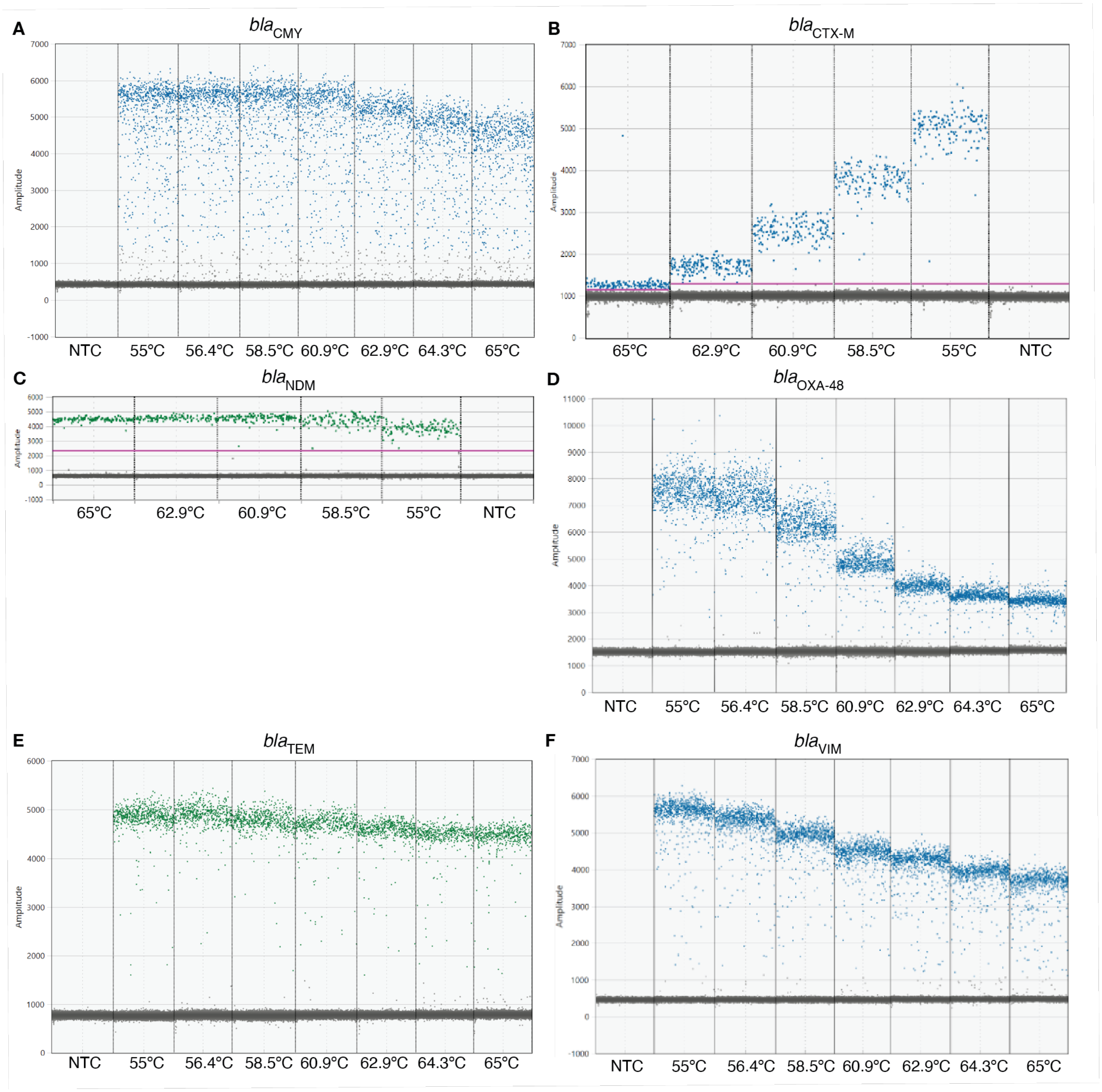


**Figure S9. Temperature gradient test results for ARG assays.** Screenshots for droplet digital PCR 1D plot of temperature gradient testing for **A)** *bla*$_{CMY}$, **B)** *bla*$_{CTX\text{-}M}$, **C)** *bla*$_{NDM}$, **D)** *bla*$_{OXA\text{-}48}$, **E)** *bla*$_{TEM}$, **F)** *bla*$_{VIM}$, **G)** *mcr-1*, **H)** *mecA*, **I)** *tetW*, and **J)** *vanA*. Temperature gradient testing was done with gblocks of target sequences (sequences provided in the SI), except for *bla*$_{NDM}$ and *bla*$_{CTX\text{-}M}$, which were done with 1:10 and 1:100 diluted wastewater respectively. Probes were either FAM (blue dots in the figure) or HEX (green dots in the figure). The pink line shows the automatic threshold by QX Manager. *bla*$_{KPC}$ was added later as the last target so instead of a temperature gradient testing, 58°C was tested to confirm that the assay performs well at the temperature set for other assays.

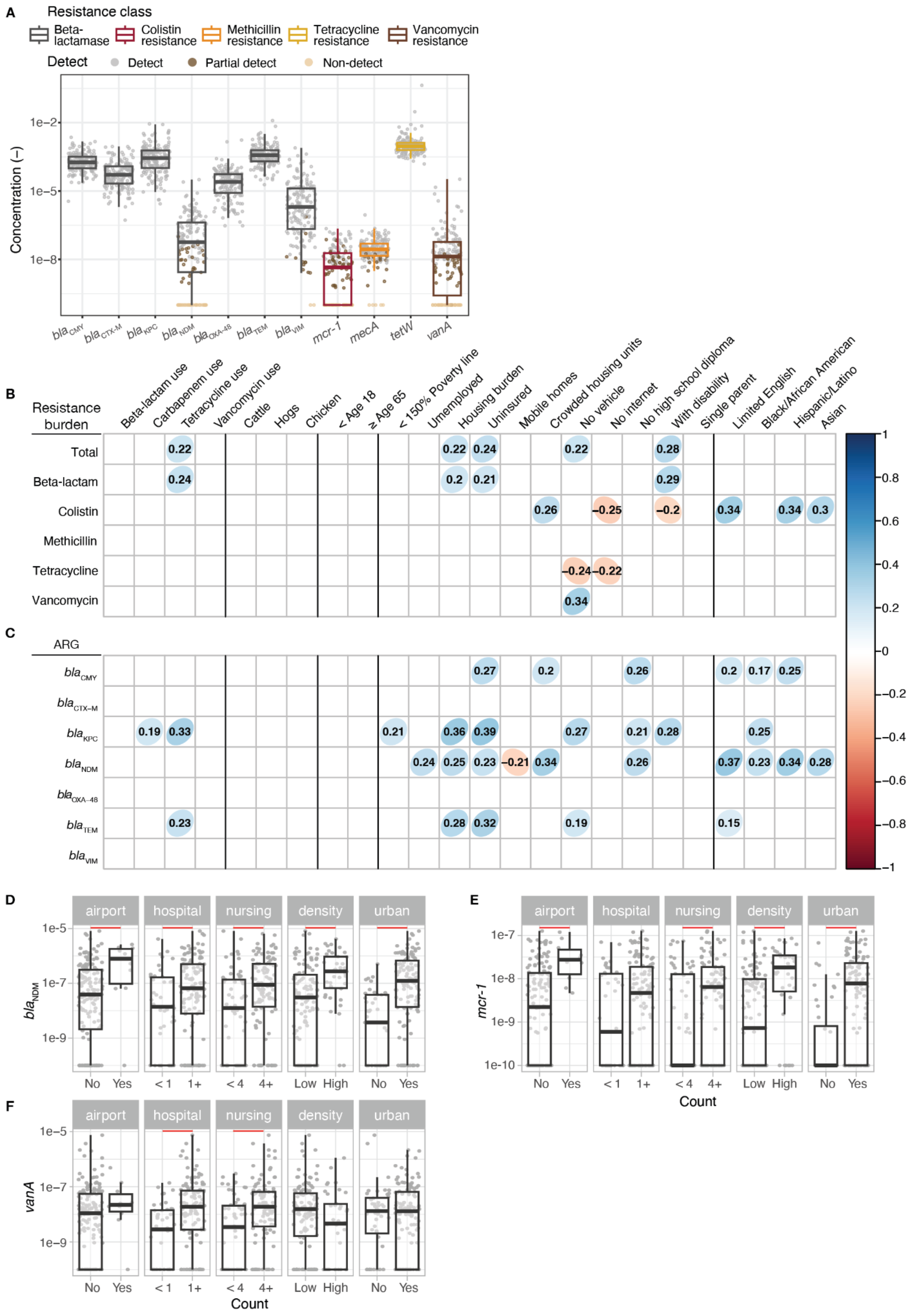
A
Resistance class
Beta-lactamase
Colistin resistance
Methicillin resistance
Tetracycline resistance
Vancomycin resistance
Detect
Detect
Partial detect
Non-detect
Concentration (–)
1e−2
1e−5
1e−8
blaCMY
blaCTX-M
blaKPC
blaNDM
blaOXA-48
blaTEM
blaVIM
mcr-1
mecA
tetW
vanA
B
Resistance burden
Beta-lactam use
Carbapenem use
Tetracycline use
Vancomycin use
Cattle
Hogs
Chicken
< Age 18
≥ Age 65
< 150% Poverty line
Unemployed
Housing burden
Uninsured
Mobile homes
Crowded housing units
No vehicle
No internet
No high school diploma
With disability
Single parent
Limited English
Black/African American
Hispanic/Latino
Asian
Total
Beta-lactam
Colistin
Methicillin
Tetracycline
Vancomycin
C
ARG
D
E
F
airport
hospital
nursing
density
urban
No
Yes
< 1
1+
< 4
4+
Low
High
Count
1e−5
1e−7
1e−9
1e-10
mcr-1
vanA

**Fig S10. Analysis using substitution of 0 for ND instead of half theoretical detection limit.** **A)** ARG concentration normalized by 16S rRNA gene for all sample sites. **B)** antibiotic resistance burden score and **C)** beta-lactamase gene concentrations measured in wastewater and normalized by 16S rRNA gene ($p < 0.05$ with Benjamin-Hochberg correction applied). Bivariate analysis of points of interest, population density, and urbancity for **D)** $bla_{NDM}$, **E)** *mcr-1*, and **F)** *vanA* concentration measured in wastewater normalized by 16S rRNA. Each symbol in panels A, D-F represents a wastewater treatment plant. 1e-10 was added to all values to show ND on a log-scale plot. The median is shown by the line inside the box with the 25th and 75th percentile represented by the lower and upper boundary of the box. Bottom and top whiskers show 1.5 x interquartile range. Red lines indicate a significant pairwise difference between regions measured by the Conover-Iman post-hoc test with Benjamini-Hochberg correction applied ($p < 0.05$).

**Table S1**. **Summary of samples collected per census region.**

| | All | Midwest | Northeast | South | West |
|---|---|---|---|---|---|
| **# of WWTPs** | 163 | 32 | 24 | 48 | 59 |
| **Total # of samples** | 443 | 92 | 59 | 130 | 162 |
| **Mean # of samples per WWTP** | 2.7 | 2.9 | 2.5 | 2.7 | 2.7 |

**Table S2. Statistics for Wilcoxon post-hoc test in Figure 2.**

| | Test statistics | Confidence interval | Effect size | Degrees of freedom | Adjusted p-values |
|---|---|---|---|---|---|
| Total burden - hospital | 1432 | (-1.6, -0.27) | 0.23 | 142 | 0.017 |
| Total burden - nursing | 1503 | (-1.8, -0.61) | 0.32 | 142 | 0.0007 |
| Colistin burden - airport | 272 | (-0.84, -0.17) | 0.27 | 155 | 0.002 |
| Colistin burden - density | 1204 | (-0.43, -0.10) | 0.31 | 155 | 2.6e-4 |
| Colistin burden - urban | 1352 | (-0.23, -0.08) | 0.35 | 155 | 6.0e-5 |
| $bla_{NDM}$ - airport | 354 | (-2.6e-6, 8.1e-6) | 0.22 | 160 | 0.005 |
| $bla_{NDM}$ - hospital | 1969 | (-8.1e-6, 4.1e-6) | 0.18 | 160 | 0.022 |
| $bla_{NDM}$ - nursing | 2096 | (-6.4e-6, 8.1e-6) | 0.25 | 160 | 0.001 |
| $bla_{NDM}$ - density | 1329 | (-4.1e-6, 8.1e-6) | 0.32 | 160 | 6.1e-5 |
| $bla_{NDM}$ - urban | 1056 | (-8.1e-6, 5.0e-7) | 0.45 | 160 | 1.5e-8 |

**Table S3**. **Assays used in this study and expected concentration (high vs low) in wastewater samples from the U.S. based on preliminary testing of seven wastewater samples.**

| Target | Sequences (5' - 3') | Expected Conc. (dilution) | Final Fluorophore | Amplicon Length | Ref. |
|---|---|---|---|---|---|
| *bla*$_{CMY}$ | F: AGACGTTTAACGGCGTGTTG<br>R: TAAGTGCAGCAGGCGGATAC<br>P: TATCGCCCGCGGCGAAAT | High (1:100) | Cy5 | 128 | [1] |
| *bla*$_{CTX-M}$ | F: CCGTCACGCTGTTRTTAGGA<br>R: AATGCCACMCCCAGYCKKCC<br>P: CAGCAAAAACTTGCCGRATT | High (1:100) | ATTO590 | 109 | [2] |
| *bla*$_{KPC}$ | F: GGCCGCCGTGCAATAC<br>R: GCCGCCCAACTCCTTCA<br>P:TGATAACGCCGCCGCCAATTTGT | High (1:100) | Cy5.5 | 61 | [3] |
| *mecA* | F: CATTGATCGCAACGTTCAATTTAAT<br>R: TGGTCTTTCTGCATTCCTGGA<br>P: CTATGATCCCAATCTAACTTCCACATACC | Low (no dilution) | ATTO590 | 99 | [4] |
| *bla*$_{NDM}$ | F: ATATCACCGTTGGGATCGAC<br>R: TAGTGCTCAGTGTCGGCATC<br>P: AAGGACAGCAAGGCCAAGTCG | Low (no dilution) | FAM | 102 | [5] |
| *bla*$_{OXA-48}$ | F: ACGGGCGAACCAAGCAT<br>R: GCGATCAAGCTATTGGGAATTT<br>P: TTACCCGCATCTACC | High (1:100) | HEX | 60 | [6] |
| *bla*$_{TEM}$ | F: GCATCTTACGGATGGCATGA<br>R: GTCCTCCGATCGTTGTCAGAA<br>P: CAGTGCTGCCATAACCATGAGTGA | High (1:100) | FAM | 100 | [7] |
| *bla*$_{VIM}$ | F: TSTACCCRTCCAATGGTCTC<br>R: AGAAGKGCCRCTGTGTTTTT<br>P:TGTCCGTGATGGYGATGAGTTG | Low (no dilution) | ROX | 91 | [2] |
| *mcr-1* | F: CATCGCGGACAATCTCGG<br>R: AAATCAACACAGGCTTTAGCAC<br>P: AACAGCGTGGTGATCAGTAGCAT | Low (no dilution) | HEX | 116 | [8] |
| *tetW* | F: GCAGAGCGTGGTTCAGTCT<br>R: GACACCGTCTGCTTGATGATAAT<br>P: TTCGGGATAAGCTCTCCGCCGA | High (1:100) | ROX | 66 | [9] |

| | | | | | |
|---|---|---|---|---|---|
| *vanA* | F: ATCAACCATGTTGATGTAGC<br>R: AAGGGATACCGGACAATTCA<br>P: TCCATCTTCACCTGACTTGCCA | Low (no dilution) | Cy5.5 | 94 | [10] |
| 16S rRNA | F: CGGTGAATACGTTCYCGG<br>R: GGWTACCTTGTTACGACTT | (1:50,000) | EvaGreen | | [11] |

All primers and probes ordered from IDT DNA (Maryland, USA).

**Table S4**. **Samples provided from each WWTP.** Tables of Locations for AMR Manuscript

**Table S5. Mean and standard deviation of the total number of copies of target per reaction for each target**

| **Target** | **n** | **mean** | **sd** |
|---|---|---|---|
| *bla*$_{CMY}$ | 445 | 0.0451 | 0.0607 |
| *bla*$_{CTX-M}$ | 445 | 0.0175 | 0.0328 |
| *bla*$_{KPC}$ | 445 | 0.0766 | 0.118 |
| *mecA* | 444 | 0.000735 | 0.00112 |
| *bla*$_{NDM}$ | 445 | 0.0106 | 0.05 |
| *bla*$_{OXA-48}$ | 442 | 0.00733 | 0.0184 |
| *bla*$_{TEM}$ | 445 | 0.0844 | 0.102 |
| *bla*$_{VIM}$ | 445 | 0.128 | 0.214 |
| *mcr-1* | 442 | 0.000260 | 0.000474 |
| *tetW* | 445 | 0.183 | 0.169 |
| *vanA* | 445 | 0.00866 | 0.072 |
| 16S rRNA | 439 | 0.321 | 0.215 |

Copies per target calculated by dividing the number of positive droplets by the number of accepted droplets. Note that mean and sd were calculated using all samples, not just those with detectable target.

gBlock sequences for assays

Assays were initially tested with gBlocks containing the target region and surrounding bases for each ARG assay. Maximum of three target regions were combined to create one gBlock as shown below, separated by a cluster of thymines.

For *bla*$_{TEM}$, *bla*$_{NDM}$ (contains sequences for *bla*$_{CTX-M}$ corresponding to an assay not used in this study):

AGAGACACCACCACGCCGCGGGCGATGGCGCAGACGTTGCGTCAGCTTACGCTGGGTCA
TGCGCTGGGCGAAACCCAGCGGGCGCAGTTGGTGACGTGGCTCAAAGGCAATACGACCG
GCGCAGCCAGCATTCGGGCCGGCTtttttACTCACCAGTCACAGAAAAGCATCTTACGGATGG
CATGACAGTAAGAGAATTATGCAGTGCTGCCATAACCATGAGTGATAACACTGCGGCCAAC
TTACTTCTGACAACGATCGGAGGACCGAAGGAGCTAACCGtttttTCGATACCGCCTGGACCG
ATGACCAGACCGCCCAGATCCTCAACTGGATCAAGCAGGAGATCAACCTGCCGGTCGCGC
TGGCGGTGGTGACTCACGCGCATCgcggccgc

For *bla*$_{CMY}$*, tetW, vanA*:

GGTCGGTCAGTAAGACGTTTAACGGCGTGTTGGGCGGCGACGCTATCGCCCGCGGCGAA
ATTAAGCTCAGCGATCCGGTCACGAAATACTGGCCAGAACTGACAGGCAAACAGTGGCGG
GGTATCAGCCTGCTGCACTTAGCCACCTATACAGCGtttttTGGCGTTGATTTGCAGAGCGTG
GTTCAGTCTGTTCGGGATAAGCTCTCCGCCGATATTATCATCAAGCAGACGGTGTCGCTGT
CCCCGGtttttATGCACGGATTACTTGTTAAAAAGAACCATGAATATGAAATCAACCATGTTGAT
GTAGCATTTTCAGCTTTGCATGGCAAGTCAGGTGAAGATGGATCCATACAAGGTCTGTTTG
AATTGTCCGGTATCCCTTTTGTAGGCTGCGATATTCAAAGCTCAGCAATTTGTATGGACAAg
cggccgc

For *mecA* (contains sequences for *mcr-1* and *bla*$_{SHV}$ corresponding to assays not used in this study):

TATCCCATCGCGGACAATCTCGGCTTTGTGCTGACGATCGCTGTCGTGCTCTTTGGCGCG
ATGCTACTGATCACCACGCTGTTATCATCGTATCGCTATGTGCTAAAGCCTGTGTTGATTTT
GCTATTAATCATGGGCGCGGTGACCAGTTATTTTACTGACACTTATGGCACtttttCTGGCGCG
CCGATGAACGCTTTCCCATGATGAGCACCTTTAAAGTAGTGCTCTGCGGCGCAGTGCTGG
CGCGGGTGGATGCCGGTGACGAACAGCTGGAGCGAAAGATCCACTATCGCCAGCAGGAT
CTGGTGGACTACTCGCCGGTCtttttTGCTCAATATAAAATTAAAACAAACTACGGTAACATTGA
TCGCAACGTTCAATTTAATTTTGTTAAAGAAGATGGTATGTGGAAGTTAGATTGGGATCATA
GCGTCATTATTCCAGGAATGCAGAAAGACCAAAGCATACATATTGAAAATTTAAAATgcggccg
c

For *bla*$_{OXA-48}$, *bla*$_{VIM}$ (contains sequence for *bla*$_{IMP}$ corresponding to an assay not used in this study):

CAAGGATTTACCAATAATCTTAAACGGGCGAACCAAGCATTTTTACCCGCATCTACCTTTAA
AATTCCCAATAGCTTGATCGCCCTCGATTTGGGCGTGGTTAAGtttttGCAGTCGTTTGATGGC
GCGGTCTACCCGTCCAATGGTCTCATTGTCCGTGATGGTGATGAGTTGCTTTTGATTGATA
CAGCGTGGGGTGCGAAAAACACAGCGGCACTTCTCGCGGAGATTGAAAAGCAAATTtttttTC
CAGGGCACACTCCAGATAACGTAGTGGTTTGGCTACCTGAAAATAGAGTTTTGTTCGGTGG

TTGTTTTGTTAAACCGTACGGTCTTGGTAATTTGGGTGACGCAAATTTAGAAGCTTGGCCAA
AGTCCGCCAAATTATTAATGTCCgcggccgc

For *mcr-1* (contains sequences for $bla_{SHV}$ and $bla_{IMP}$ corresponding to assays not used in this study):
CCCGAATAACAAAGCAGAGCGCATTGTAGTGATTTATCTGCGGGATACCCCGGCGAGCAT
GGCCGAGCGAAATCAGCAAATCGCCGGGATCGGCGCGGCGCTGATCGAGCACTGGCAAC
GCTAAtttttCAGCACGGGCGGAATAGAGTGGCTTAATTCTCGATCTATCCCCACGTATGCATC
TGAATTAACAAATGAACTGCTTAAAAAAGACGGTAAGGTTCAAGCCACAAATTCATTTAGCG
GAGTTAACTATTGGCTAGTTAAAAATAAAATTGAAGTTTTTTATCCAGGCCCGGGACACACT
CCAGATAACGTAGTGGTTTGGTTGCCTGAAAGGtttttGCCAAACCTATCCCATCGCGGACAAT
CTCGGCTTTGTGCTGACGATCGCTGTCGTGCTCTTTGGCGCGATGCTACTGATCACCACG
CTGTTATCATCGTATCGCTATGTGCTAAAGCCTGTGTTGATTTTGCTATTAATCATGGgcggcc
gc

For $bla_{KPC}$ (contains sequence for *vanB* corresponding to an assay not used in this study):
GACGGTGGCGGAGCTGTCCGCGGCCGCCGTGCAATACAGTGATAACGCCGCCGCCAATT
TGTTGCTGAAGGAGTTGGGCGGCCCGGCCGGGCTGACGGCCtttttGGGGAACGAGGATGA
TTTGATTGTCGGCGAAGTGGATCAAATCCGGCTGAGCCACGGTATCTTCCGCATCCATCAG
GAAAACGAGCCGGAAAAAGGCgcggccgc

#### Preparation of secondary data

***Demographic and socioeconomic factors.*** The 2022 Social Vulnerability Index (SVI) from the US CDC includes various demographic and socioeconomic factors from the 2022 5-year American Community Survey (ACS) that relate to a community's social vulnerability or ability to respond to external hazards.[12] For each sewershed, we approximated the value of selected variables used in the SVI as a proportion using the relevant ACS variables obtained at the census tract resolution (**Table S6**).[13] Census tract and sewershed boundaries did not align, so we first aggregated ACS variable counts across all census tracts intersecting a sewershed to estimate counts for each sewershed. Specifically, we used the Tabulate Intersection geoprocessing tool in ArcGIS Pro (version 3.1.1) to determine the area proportion $p$ of each intersecting census tract $n$ in the sewershed.[14] Next, we adjusted the census tract-level count of each ACS variable based on $p$ (**Equation 1**). Then we summed the adjusted counts across all $N$ census tracts intersecting the sewershed to determine the sewershed-level count for each ACS variable (**Equation 2**). Using sewershed-level counts of each ACS variable, we calculated the sewershed-level value of each SVI variable as a proportion as described in **Table S6**. The distribution of each proportion among sewersheds in the study is shown in **Figure S11**.

**Equation 1**. $count\_adjusted_n = count_n \times p_n$
**Equation 2**. $count\_sewershed \; = \sum_{n=1}^{N} \quad count\_adjusted_n$

**Table S6. Selected main and adjunct variables in the Social Vulnerability Index (SVI) calculated as proportions using relevant American Community Survey (ACS) variables**

| Proportion | Description | Calculation using ACS variables[a] |
|---|---|---|
| < 150% Poverty line | Proportion of the population for whom poverty status is determined below the 150% poverty line | S1701_C01_040E / S1701_C01_001E |
| Unemployed | Proportion of the civilian labor force age 16+ years that is unemployed | DP03_0005E / DP03_0003E |
| Housing burden | Proportion of occupied housing units that are housing cost-burdened with an annual income <$75K | (S2503_C01_028E + S2503_C01_032E + S2503_C01_036E + S2503_C01_040E) / S2503_C01_001E |
| No high school diploma | Proportion the population age 25+ years with no high school diploma | B06009_002E / B06009_001E |
| Uninsured | Proportion of the civilian noninstitutionalized population that is uninsured | S2701_C04_001E / S2701_C01_001E |
| ≥ Age 65 | Proportion of the population age 65+ years | S0101_C01_030E / S0601_C01_001E |
| < Age 18 | Proportion of the population age 17 years and younger | DP05_0019E / S0601_C01_001E |

| | | |
|---|---|---|
| With disability | Proportion of the civilian noninstitutionalized population with a disability | DP02_0072E / S2701_C01_001E |
| Single parent | Proportion of single-parent households with children <18 years | (DP02_0007E + DP02_0011E) / (DP02_0006E + DP02_0010E) |
| Limited English | Proportion of persons age 5+ years who speak English "less than well" | (B16005_007E + B16005_008E + B16005_012E + B16005_013E + B16005_017E + B16005_018E + B16005_022E + B16005_023E + B16005_029E + B16005_030E + B16005_034E + B16005_035E + B16005_039E + B16005_040E + B16005_044E + B16005_045E) / B16005_001E |
| Black/African American | Proportion of the population that is Black/African American, not Hispanic or Latino | DP05_0080E / S0601_C01_001E |
| Hispanic/Latino | Proportion of the population this is Hispanic or Latino | DP05_0073E / S0601_C01_001E |
| Asian | Proportion of the population that is Asian, not Hispanic or Latino | DP05_0082E / S0601_C01_001E |
| Mobile homes | Proportion of housing units that are mobile homes | DP04_0014E / DP04_0001E |
| Crowded housing units | Proportion of occupied housing units with more people than rooms | (DP04_0078E + DP04_0079E) / DP04_0002E |
| No vehicle | Proportion of occupied housing units with no vehicle | DP04_0058E / DP04_0002E |
| No internet | Proportion of households with no internet | S2801_C01_019E / DP02_0001E |

[a] Variables are from the 2022 5-year American Community Survey (ACS). Census tract-level counts for each variable were first adjusted and summed by sewershed to obtain sewershed-level counts.

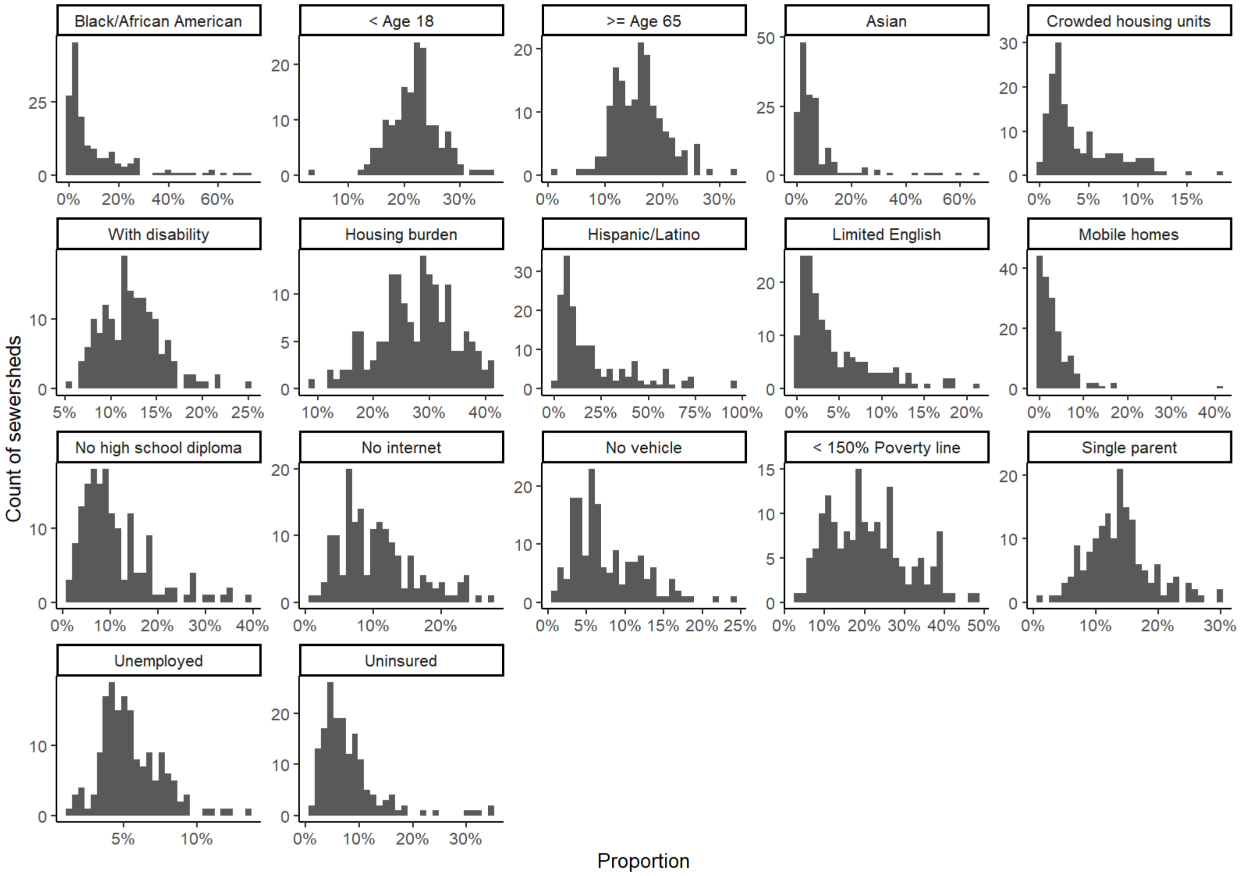


**Figure S11**. **Distribution of each variable in Table S6 among sewersheds.**

***Urbanicity and population density.*** We used 2020 urban-rural designations from the US Census Bureau to determine the extent to which sewersheds are classified as urban.[15] For each sewershed, we used the Tabulate Intersection geoprocessing tool in ArcGIS Pro to determine the area proportion of the sewershed that intersects any urban area. We assigned a value of 0 if a sewershed did not intersect any urban area. The distribution of the urban proportion among sewersheds in this study is shown in **Figure S12**. Sewersheds with an urban area proportion ≥ 50% were classified as urban and sewersheds with an urban area proportion < 50% were classified as non-urban for data analysis. To determine population density, we obtained the population of each county from the 2022 5-year ACS (subject table S0101) and the land area in square meters of each county from the 2022 US Census Bureau county boundaries (5-meter resolution).[13,14] We calculated the population density in square kilometers of each county by dividing the population by the land area and multiplying by $1000^2$. We assigned the population density of each sewershed as the population density of its predominant county (i.e., the county that the majority of the sewershed is located in). We determined the predominant county of each sewershed using the Tabulate Intersection geoprocessing tool in ArcGIS Pro and 2022 county boundaries (5-meter resolution) from the US Census Bureau.[14] The distribution of population density among sewersheds in this study is shown in **Figure S13**. Population density was stratified by 617 per $km^2$, the average number of people in cities in the U.S., for data analysis.

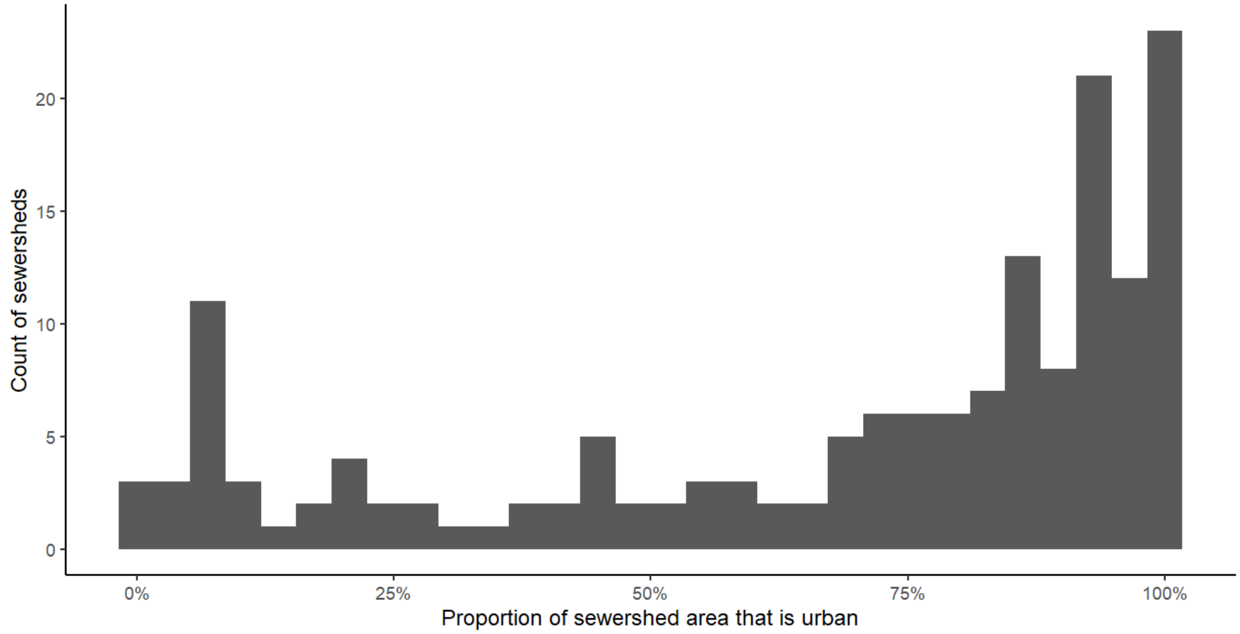


**Figure S12**. **Distribution of urbanicity among sewersheds.**

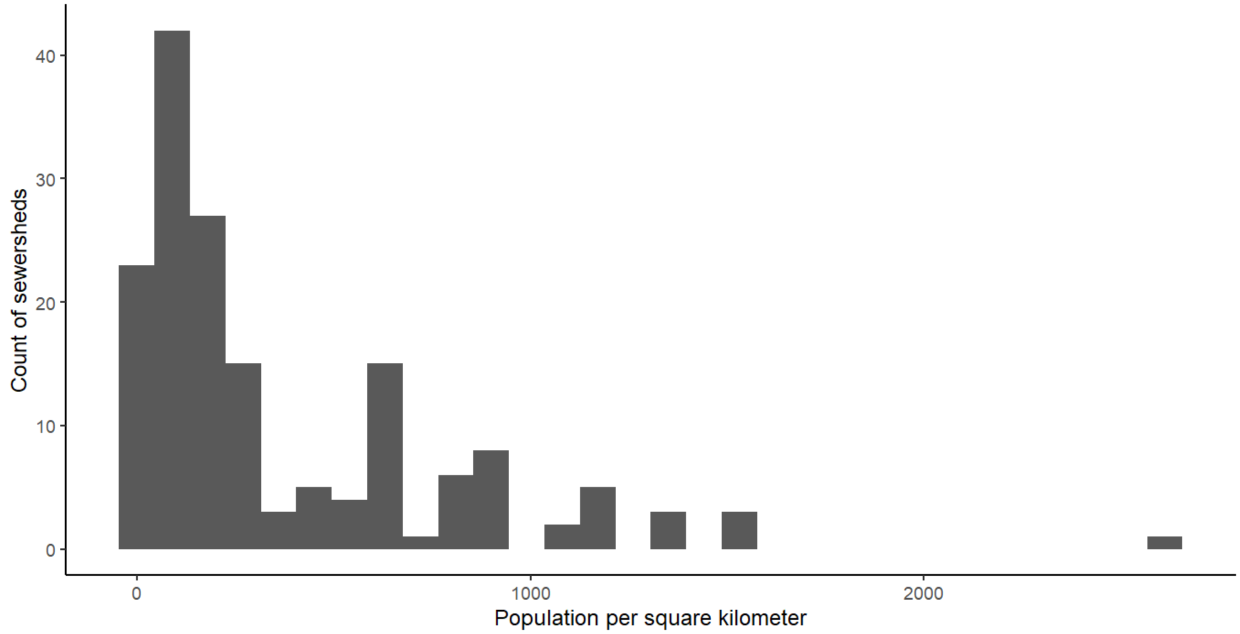


**Figure S13**. **Distribution of population density among sewersheds.**

***Points of interest.*** We obtained the locations of major airports (defined as usage category "1,000,000 or more") from Esri (source: Federal Aviation Administration's National Airspace System Resource Aeronautical Data Product).[16] We obtained the locations of hospitals and nursing homes from the US Department of Homeland Security's Homeland Infrastructure Foundation-Level Data database.[17] We used the Tabulate Intersection geoprocessing tool in ArcGIS Pro to determine the number of airports, hospitals, and nursing homes in each sewershed. For hospitals and nursing homes, we omitted any locations with a non-open status prior to using the Tabulate Intersection tool. **Figure S14** shows the distribution of airports, hospitals, and nursing homes among sewersheds in this study. We could not weight hospitals and nursing homes by size due to not all sites reporting a valid value for population or beds. For data analysis, airports and hospitals were split into groups based on the presence of the facilities, and the number of nursing homes were split by the national median number of nursing homes (median: 4).

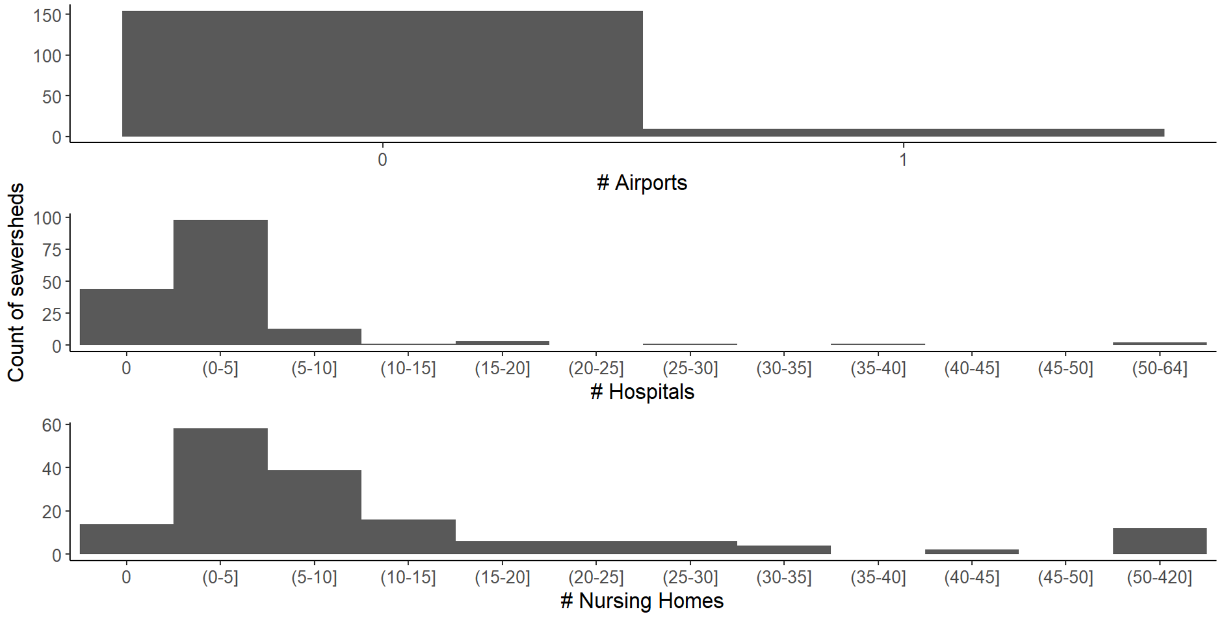


**Figure S14**. **Distribution of points of interest among sewersheds.**

***Agricultural activity.*** Using the US Department of Agriculture (USDA) National Agricultural Statistics Service (NASS), we obtained 2022 inventory numbers for cattle, chickens, and hogs at the county resolution (**Table S7**).[18] For each sewershed, we assigned the inventory numbers of its predominant county (described above) to use as an indicator of agricultural activity in the sewershed. If inventory values were not reported or withheld to disclose numbers for individual operations, we assumed a value of zero. The Newark, NJ sewershed had no reported data for its predominant county, so we assigned inventory numbers from its second most predominant county to use for the analysis. The Census Bureau provides boundaries for county equivalents in Connecticut rather than counties which are used in the USDA NASS dataset. We assumed the Western Planning Region county equivalent is most similar to Fairfield county to determine the predominant county for the Stamford, CT sewershed. For chickens, we summed over all data items (broilers, layers, pullets, roosters). The distribution of cattle, chickens, and hogs among sewersheds in the study is shown in **Figure S15**.

**Table S7. US Department of Agriculture National Agricultural Statistics Service Selections**

| | |
|---|---|
| **Program** | **Census** |

| | |
|---|---|
| Sector | Animals & Products |
| Group | Poultry; Livestock |
| Commodity | Cattle; Chickens; Hogs |
| Category | Inventory |
| Data Item | Cattle, Incl Calves - Inventory<br>Chickens, Broilers - Inventory<br>Chickens, Layers - Inventory<br>Chickens, Pullets, Replacement - Inventory<br>Chickens, Roosters - Inventory<br>Hogs - Inventory |
| Domain | Total |
| Geographic Level | County |
| State | All |
| Year | 2022 |
| Period Type | Point in Time |
| Period | End of Dec |

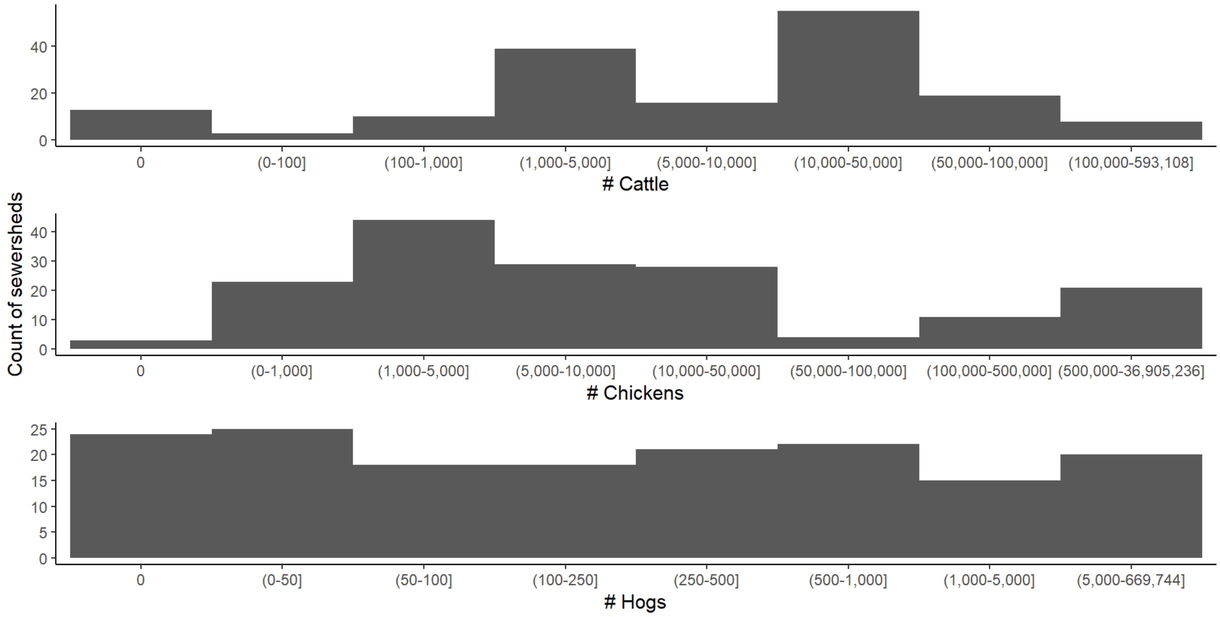


Figures S15. Distribution of agriculture animal inventory numbers among sewersheds.

Epic Cosmos data. All encounters in the Epic Cosmos dataset were temporally limited to encounters from June 1, 2023–June 1, 2024. Encounters for sampled sewersheds were further geographically limited to the 116 counties representing the predominant county serviced by wastewater treatment plants (described above). Encounters were then filtered for all encounters with beta-lactam antibiotics (penicillins, cephalosporins, and carbapenems), tetracycline, vancomycin, and colistin antibiotics dispensed. The annual numbers of encounters with these prescriptions were then divided by the total number of encounters in the analysis period for each county. Minimal colistin use was reported in the Epic dataset, so this variable was ultimately not included.

### Environmental Microbiology Minimum Information Checklist

#### Study Description

Study: [Study Name]
Date: 07-Jul-2021
Completed by: [Filled By...]

| Environmental Sampling | Sample Treatment | Sample Reduction | Nucleic Acid Extraction | Reverse Transcription | PCR Detection | Analysis |
|---|---|---|---|---|---|---|
| Described in methods section | ☐ Performed<br>No sample treatment performed | ☐ Performed<br>Centrifugation was used, as described in the methods and referenced paper | Methods described briefly in pre-analytical processing and nucleic-acid extraction section and in references. | ☐ Performed<br>Not performed | ☐ qPCR ■ dPCR<br>Methods provided including the dMIQE checklist | QA/QC criteria and analysis described in methods |

#### Control Checklist

| | Environmental Sampling | Sample Treatment | Sample Reduction | Nucleic Acid Extraction | Reverse Transcription | PCR Detection |
|---|---|---|---|---|---|---|
| Step performed | ■ | ☐ | ■ | ■ | ☐ | ■ |
| **Negative Controls** | | | | | | |
| Step has control info | ☐ | ☐ | ☐ | ☐ | ☐ | ■ |
| # control replicates | 0 | 0 | 0 | 3 | 0 | 3 |
| Control result reported | ☐ | ☐ | ☐ | ☐ | ☐ | ■ |
| Data handling reported | ☐ | ☐ | ☐ | ☐ | ☐ | ■ |
| **Positive Controls** | | | | | | |
| Control introduced | ☐ | ☐ | ☐ | ■ | ☐ | ■ |
| Internal/External | N/A | N/A | Internal | External | N/A | External |
| Independent/Parallel | N/A | N/A | Parallel | Independent | N/A | Independent |
| Step has control info | ☐ | ☐ | ☐ | ■ | ☐ | ■ |
| # control replicates | 0 | 0 | 0 | 1 | 0 | 1 |
| Control result reported | ☐ | ☐ | ☐ | ■ | ☐ | ■ |
| Data Handling reported | ☐ | ☐ | ☐ | ■ | ☐ | ■ |

#### Process Checklist

##### Environmental Sampling

- ■ Sampling Procedure
- ■ Number of samples
- ■ Sample amount, mean, range
- ■ Sampling locations, dates, times

##### Sample Treatment

- ☐ Performed
- ☐ Treatment procedure
- ☐ Reagents

##### Sample Reduction

- ■ Performed
- ■ Reduction procedure
- ■ Reagents
- ☐ Concentration Factor

##### Nucleic Acid Extraction

- ■ Extraction procedure
- ■ Amount extracted, amount obtained
- ■ Extract storage conditions

##### qPCR or dPCR

- ■ Target gene name, amplicon length
- ■ Thermocycling temperatures and times
- ■ Master mix: composition, vendors, concentrations
- ■ Additives: vendors, concentrations
- ■ Template amount added, pre-treatment (if any)
- ■ Primers: sequences, concentrations, vendors, references
- ■ Amplicon confirmation method (probe, melt curve, etc)
- ■ Probe sequence, concentration, vendor, reference
- ■ Instrumentation
- ☐ Equivalent volume of sample analyzed by PCR
- ■ Inhibition assessment procedure
- ■ Inhibition control description (if used)
- ■ Number samples tested and found inhibited

##### Reverse Transcription

- ☐ Performed
- ☐ One or two step
- ☐ cDNA storage conditions (if two step)
- ☐ Reaction temperatures and times
- ☐ Reaction reagents and concentrations
- ☐ Priming method
- ☐ Reaction volume, added template amount
- ☐ Inhibition assessment procedure
- ☐ Inhibition control description (if used)
- ☐ Number samples tested and found inhibited

##### Analysis – dPCR

- ■ Threshold settings
- ■ Technical replicates, number, well merging
- ■ Partitions measured, number, mean, variance
- ■ Partition volume
- ■ Target copies per partition, mean, variance
- ■ Program used for dPCR analysis
- ■ Explanation of control results, example plots

##### Analysis – qPCR

- ☐ Method for handling failed negative controls
- ☐ Technical replicates, number, calculations
- ☐ Calibration standards: description and source
- ☐ Method of quantifying standards
- ☐ Calibration curve slope
- ☐ Calibration curve R2
- ☐ Lowest standard measured or 95% LOD
- ☐ Cq value determination method

#### References

1. Schmidt, G. V. *et al.* Sampling and Pooling Methods for Capturing Herd Level Antibiotic Resistance in Swine Feces using qPCR and CFU Approaches. *PLOS ONE* **10**, e0131672 (2015).
2. Pholwat, S. *et al.* Genotypic antimicrobial resistance assays for use on E. coli isolates and stool specimens. *PLOS ONE* **14**, e0216747 (2019).
3. Garcia, L. S. *Clinical Microbiology Procedures Handbook, 3rd Ed.* (ASM Press, 2010).
4. Böckelmann, U. *et al.* Quantitative PCR Monitoring of Antibiotic Resistance Genes and Bacterial Pathogens in Three European Artificial Groundwater Recharge Systems. *Appl. Environ. Microbiol.* **75**, 154–163 (2009).
5. Chavda, K. D. *et al.* Evaluation of a Multiplex PCR Assay To Rapidly Detect Enterobacteriaceae with a Broad Range of β-Lactamases Directly from Perianal Swabs. *Antimicrob. Agents Chemother.* **60**, 6957–6961 (2016).
6. Lutgring, J. D. *et al.* Phenotypic and Genotypic Characterization of *Enterobacteriaceae* Producing Oxacillinase-48–Like Carbapenemases, United States. *Emerg. Infect. Dis.* **24**, 700–709 (2018).
7. Roschanski, N., Fischer, J., Guerra, B. & Roesler, U. Development of a Multiplex Real-Time PCR for the Rapid Detection of the Predominant Beta-Lactamase Genes CTX-M, SHV, TEM and CIT-Type AmpCs in Enterobacteriaceae. *PLoS ONE* **9**, e100956 (2014).
8. Yang, D. *et al.* The Occurrence of the Colistin Resistance Gene mcr-1 in the Haihe River (China). *Int. J. Environ. Res. Public. Health* **14**, 576 (2017).
9. Smith, M. S. *et al.* Quantification of Tetracycline Resistance Genes in Feedlot Lagoons by Real-Time PCR. *Appl. Environ. Microbiol.* **70**, 7372–7377 (2004).
10. Farivar, T. N. *et al.* Development and evaluation of a Quadruplex Taq Man real-time PCR assay for simultaneous detection of clinical isolates of Enterococcus faecalis, Enterococcus

faecium and their vanA and vanB genotypes. *Iran. J. Microbiol.* **6**, 335–340 (2014).

11. Suzuki, M. T., Taylor, L. T. & DeLong, E. F. Quantitative Analysis of Small-Subunit rRNA Genes in Mixed Microbial Populations via 5′-Nuclease Assays. *Appl. Environ. Microbiol.* **66**, 4605–4614 (2000).
12. U.S. Centers for Disease Control and Prevention. Social Vulnerability Index 2022. https://www.atsdr.cdc.gov/place-health/php/svi/svi-data-documentation-download.html?CDC_AAref_Val=https://www.atsdr.cdc.gov/placeandhealth/svi/data_documentation_download.html.
13. U.S. Census Bureau. 2022 5-Year American Community Survey (ACS). https://www.census.gov/programs-surveys/acs.
14. U.S. Census Bureau. Cartographic Boundary Files. https://www.census.gov/geographies/mapping-files/time-series/geo/cartographic-boundary.html.
15. U.S. Census Bureau. 2020 Census Urban-Rural Classification. https://www.census.gov/programs-surveys/geography/guidance/geo-areas/urban-rural.html (2024).
16. Esri Data and Maps. USA Airports 1,000,000 or more. https://hub.arcgis.com/datasets/esri::usa-airports/about?layer=1.
17. US Department of Homeland Security. Homeland Infrastructure Foundation-Level Data. https://hifld-geoplatform.hub.arcgis.com/ (2024).
18. U.S. Department of Agriculture. 2022 National Agricultural Statistics Service Quick Stats. https://quickstats.nass.usda.gov/.
